# Association of 24-hour activity patterns with risk of Alzheimer’s disease, Parkinson’s disease, and cognitive decline

**DOI:** 10.1101/2023.03.29.23287916

**Authors:** Joseph R. Winer, Renske Lok, Lara Weed, Zihuai He, Kathleen L. Poston, Elizabeth C. Mormino, Jamie M. Zeitzer

## Abstract

Sleep-wake regulating circuits are affected during prodromal stages in the pathological progression of both Alzheimer’s disease (AD) and Parkinson’s disease (PD). Assessment of 24-hour rhythm impairment may serve as an early indicator of disease and cognitive decline. Our objective was to determine whether objective markers of 24-hour activity are associated with subsequent development of AD, PD, and cognitive decline. This longitudinal study obtained UK Biobank data from 82,829 individuals with valid accelerometer data (collected from June 2013 to January 2016) out of 103,671 eligible adults aged 40 to 79 years with a mean (±SD) follow-up of 6.8 (±0.9) years. AD and PD diagnoses were ascertained through September 2021, with data analysis conducted March to November 2022. The outcomes were accelerometer-derived measures of 24-hour activity (derived by cosinor, nonparametric, and functional principal component methods), incident AD and PD diagnosis (obtained through hospitalization or primary care records), longitudinal tests of cognitive function, and demographic characteristics. 82,829 participants consisted of 46,683 women (56%) and 36,146 men (44%) with a mean (±SD) age of 62.0 (±7.8) years at the time of actigraphy data collection. During the follow-up period, 191 individuals converted to AD (0.2%) and 266 to PD (0.3%). After adjusting for covariates, interdaily stability, a measure of regularity, (hazard ratio [HR] per SD increase 1.24, 95% confidence interval [CI] 1.04-1.47), diurnal amplitude (HR 0.77, CI 0.64-0.93), mesor (mean activity; HR 0.73, CI 0.56-0.95), and activity during most active 10 hours (HR 0.73, CI 0.59-0.91), were each associated with an increased risk of AD. Diurnal amplitude (HR 0.28, CI 0.23-0.34), mesor (HR 0.12, CI 0.10-0.16), activity during least active 5 hours (HR 0.24, CI 0.08-0.68), and activity during most active 10 hours (HR 0.20, CI 0.16-0.25), were associated with an increased risk of PD. Several measures were additionally predictive of longitudinal cognitive test performance. In this community-based longitudinal study, objective measures of 24-hour activity were associated with elevated risk of AD, PD, and accelerated cognitive decline, suggesting actigraphy-estimated 24-hour rhythm integrity may serve as a scalable early marker of neurodegenerative disease.

## INTRODUCTION

Alzheimer’s disease (AD) and Parkinson’s disease (PD) are the two most prevalent neurodegenerative diseases, with numbers predicted to increase in the coming decade^1,2^. Common to both diseases is a prodromal phase during which misfolded proteins begin to spread throughout the brain, often years before clinical diagnosis^3,4^. Altered sleep and circadian rhythms, thought to result from the earliest pathology within brainstem nuclei, frequently manifest as one of the initial symptoms during this prodromal disease stage^5–7^. Thus, detection of disrupted 24-hour patterns of activity has the potential to serve as an early marker for both AD and PD.

Supporting this hypothesis, previous work using wrist-worn accelerometers has demonstrated that specific impairments in 24-hour activity patterns are associated with accelerated cognitive decline^8–12^, an increased risk of developing AD^8,9,13,14^, and an increased risk of developing PD^15^. As more older adults elect to track their daily physical activity and sleep using consumer-grade devices, attention has turned to how these patterns of activity can be leveraged in large, community-based samples. However, few studies have utilized objective assessments of 24-hour activity in large cohorts of older adults, and those that have are limited by sample sizes under 5000 with limited numbers of individuals converting to Alzheimer’s or Parkinson’s disease during the follow-up period^9,12,13,15,16^. The UK Biobank dataset thus provides a unique opportunity to explore these associations in an unprecedently large sample of individuals.

We determined whether accelerometer-derived metrics of 24-hour activity patterns are associated with longitudinal cognitive test performance and risk of incident AD and PD in a cohort of 82,829 community-dwelling older adults. By utilizing multiple approaches to 24-hour activity analysis, we sought to characterize the fragmentation and regularity of 24-hour rhythms (with nonparametric methods) as well as the robustness of activity patterns (with cosinor methods). We hypothesized that greater fragmentation and lower levels of activity would be prospectively associated with increased risk of AD, PD, and declining cognitive performance.

## METHODS

Wrist-worn triaxial accelerometry [AX3, Axivity, Newcastle upon Tyne, UK] was obtained over one week from a large community-based sample of adults living in England, Wales, and Scotland participating in the UK Biobank study, an ongoing community-based cohort study.

More than 500,000 participants aged between 40 and 69 years were recruited in 2006-2010 (full details available in Doherty et al. 2017^17^). The UK Biobank study did not exclude based on any health condition; age was the only eligibility criteria for participating in the study. No restrictions were placed on behavior during the week of accelerometer data collection. Data were downloaded March 2022.

### Participants

An overview of the inclusion process is displayed in **Supplement Figure 1**. Accelerometer data (UK Biobank data field 90001) were collected from June 2013 to January 2016 and included 103,670 total individuals. After accounting for missing and unreliable data as well as data collected during or just following daylight savings time, the remaining accelerometer data sample comprised 82,829 individuals (**Supplement Figure 1**).

All UK Biobank participants provided written informed consent. Ethical approval was obtained from the North West Multicentre Research Ethics Committee, the National Information Governance Board for Health and Social Care in England and Wales, and the Community Health Index Advisory Group in Scotland.

### Accelerometer data

#### Preprocessing

High frequency (100 Hz) accelerometer data were processed on Sherlock, a high-performance computing cluster provided by Stanford University,using the steps outlined in Weed et al. 2022^18^. In brief, data spanning 1 week of collection were down-sampled to 30 second epochs using the biobankAccelerometerAnalysis package in Python v3.6.1^17^. Non-wear time was defined as stationary episodes lasting for at least 60 minutes in which all three axes had a standard deviation of less than 13.0 mg. If present, non-wear segments were automatically imputed using the median of similar time-of-day vector magnitude and intensity distribution data points with 30-second granularity on different days of the measurement^18^. Following these preprocessing steps, we derived the following six metrics.

#### Cosinor

Cosinor analyses (fitting a cosine wave) were performed using the cosinor2 package in R^19^ and resulted in two metrics of interest: 1) mesor (rhythm adjusted mean activity) and 2) amplitude (half the difference between peak and nadir of fitted cosine wave).

#### IV-IS

Nonparametric analyses were conducted with nparACT package in R^20,21^ to derive four metrics: 1) intradaily variability (IV; fragmentation of activity within 24-hour periods), 2) interdaily stability (IS; regularity of activity across 24-hour periods), 3) activity level during the least active five hours (L5), and 4) activity level during the most active 10 hours (M10). IV values can vary from 0-2, with higher values indicating greater fragmentation. IS values can vary from 0-1, with lower values indicating lower regularity.

#### fPCA

Functional principal component analyses (fPCA) were performed using the fpca package in R^22^. Following previously established methods^22–24^, each individual’s 24-hour median accelerometer data was fit with a nine-Fourier-based function. These functions were examined with functional data analysis to determine orthogonal components that explained the most variance across individuals. The first four fPCA components (explaining >90% of the variance) were used for analysis, with individuals in a sample scored on the magnitude of each component contributing to their activity pattern. These scores were extracted and compared between disease converters and non-converters. As the results of fPCA are dependent on the specific subsets of data analyzed, we conducted two separate fPCA analyses, consisting of individuals who developed AD or PD (see below) and matched controls who did not develop AD or PD (1:1 match on age at accelerometer collection, gender, education, general health, body mass index, and Townsend Deprivation Index, using the MatchIt package in R^25^).

#### Ascertainment of Incident AD & PD

Incident AD or PD was ascertained from algorithmically defined UK Biobank variables that utilized a combination of self-report and ICD-10 codes from medical records to determine the first recorded date of a diagnosis (AD, data field 42020; PD, 42032)^26–28^. An individual would be considered to have developed AD or PD during the study period if they had an initial diagnosis of AD or PD between the time of actigraphy data collection (as early as June 2013) and September 2021. Individuals with AD (n=13) and PD (n=133) diagnoses preceding their actigraphy data collection were excluded from the AD and PD analyses, respectively.

#### Longitudinal cognitive data

Cognitive tests were designed specifically for the UK Biobank and were administered unsupervised using a computer touchscreen interface either during an in-person visit or remotely^29^. Five test scores were included from the UK Biobank cognitive battery: Trail Making Test A (numeric; 20156 and 6348) and Trail Making Test B-A (alphanumeric-numeric; 20157 and 6350), Symbol Digit Substitution Test (23323, 23324, 20159, and 20195), Numeric Memory Test (4282 and 20240), and Fluid Intelligence Test (20016 and 20191). Individuals were included in the analysis for a given test if they had (1) a test score within a year of their accelerometer data collection and (2) at least one follow-up test score. The sample available for each of the five longitudinal test scores ranged between n=6,629 and 11,030. **Supplement Table 1** outlines details for the sample and number of time points available for each test.

#### Statistical analysis

All statistical analyses were performed using R (version 4.2.1). All statistical models controlled for age at actigraphy collection, gender (31), college education (6138), baseline self-rated general health (2178), baseline body mass index (BMI; 21001), and baseline Townsend deprivation index (TDI; 189). TDI is a z-score transformed variable with scores lower than 0 indicating an area’s relative affluence and higher than 0 indicating relatively high material deprivation. Missing BMI, TDI, general health, and education data (a maximum of 1%) were imputed using the Amelia II package in R^30^. All data were otherwise complete.

Cox proportional hazard models were used to test associations between 24-hour activity rhythm metrics with incident AD and PD using the coxphf and survival R packages.For individuals who developed AD or PD, follow-up time was calculated as the interval from actigraphy data collection to AD or PD diagnosis. For individuals who did not develop AD or PD, follow-up time was the interval from actigraphy data collection to either death or the medical record censor date of 09/30/2021.

Mann-Whitney U tests were used to compare fPCA scores between converters and matched controls in the AD and PD analyses.

Longitudinal cognitive change was examined with linear mixed regression models using the lme4 R package. All linear mixed models included a random intercept for each participant, and an interaction with time for the actigraphy measure of interest and each covariate. In addition to the covariates listed above, linear mixed models included a binary variable indicating whether the baseline assessment was in-person or remote for a given individual to account for performance differences across assessment settings. Data from linear mixed models are presented as unstandardized estimates ± standard error.

## RESULTS

Participant demographics are summarized in **Table 1**. 82,829 individuals with actigraphy data were included in the study, consisting of 46,683 women (56%) and 36,146 men (44%) with a mean (±SD) age of 62.0 (±7.8) years at the time of actigraphy data collection. Correlations between the six actigraphy metrics are reported in **Supplement Table 2**.

**TABLE.** Demographics at time of actigraphy collection.

|  |  |
| --- | --- |
| **Age at baseline** | 62.0 ± 7.8 |
| **Female N (%)** | 46,683 (56) |
| **College or University degree N (%)** | 35,548 (43) |
| **General Health [1-4]** | 2.0 ± 0.7 |
| **Body mass index** | 26.7 ± 4.5 |
| **Townsend Deprivation Index** | -1.7 ± 2.8 |
| **Follow-up (years)** | 6.8 ± 0.9 |
| **Symbol Digit Substitution Score** | 20.4 ± 4.9 |
| **Trail Making Test A Duration (seconds)** | 38.0 ± 13.7 |
| **Trail Making Test B-A (seconds)** | 25.5 ± 17.9 |
| **Numeric Memory Score** | 7.04 ± 1.43 |
| **Fluid Intelligence Score** | 6.68 ± 2.03 |
| **Interdaily Stability (IS)** | 0.54 ± 0.12 |
| **Intradaily Variability (IV)** | 0.91 ± 0.24 |
| **Least Active 5 Hours (L5)** | 3.28 ± 6.41 |
| **Most Active 10 Hours (M10)** | 47.4 ± 17.1 |
| **Amplitude** | 24.4 ± 9.42 |
| **Mesor** | 28.2 ± 11.7 |

### Associations of 24-hour rhythms with risk of AD and PD

During 8.3 years (6.8 ±0.9 years) following initial actigraphic assessment, 191 individuals converted to AD (0.2%) and 266 to PD (0.3%). Survival analyses are displayed visually in **Figures 1 and 2** and summarized in **Supplement Table 3**.

**Figure 1.**
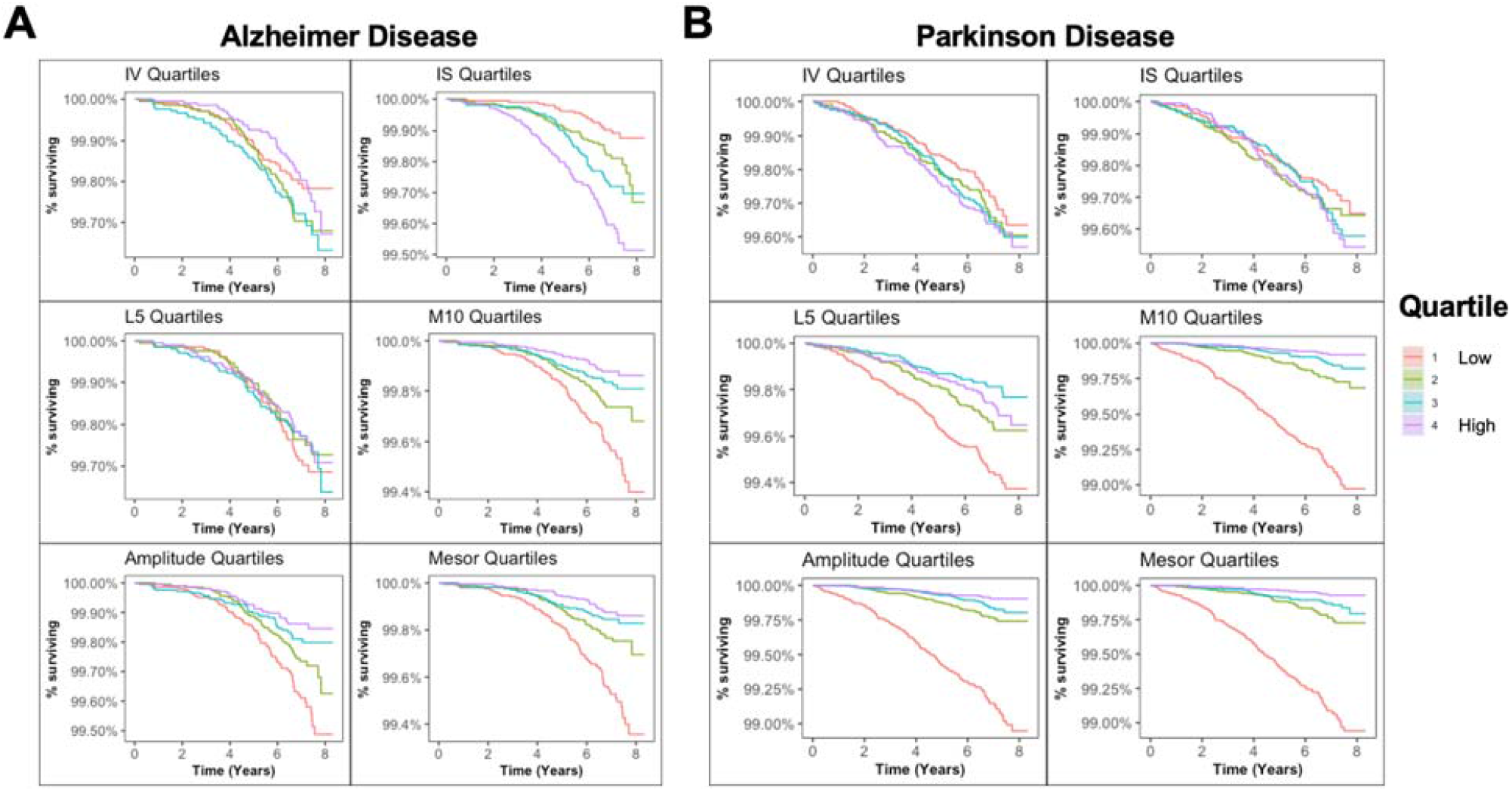
Conversion to Alzheimer’s and Parkinson’s disease by 24-hour rhythm quartiles. Survival curves for (A) Alzheimer’s disease and (B) Parkinson’s disease based on quartiles for each of the 24-hour rhythm metrics. Abbreviations: IV, intradaily variability; IS, interdaily stability, L5, least active 5 hours; M10, most active 10 hours.

**Figure 2.**
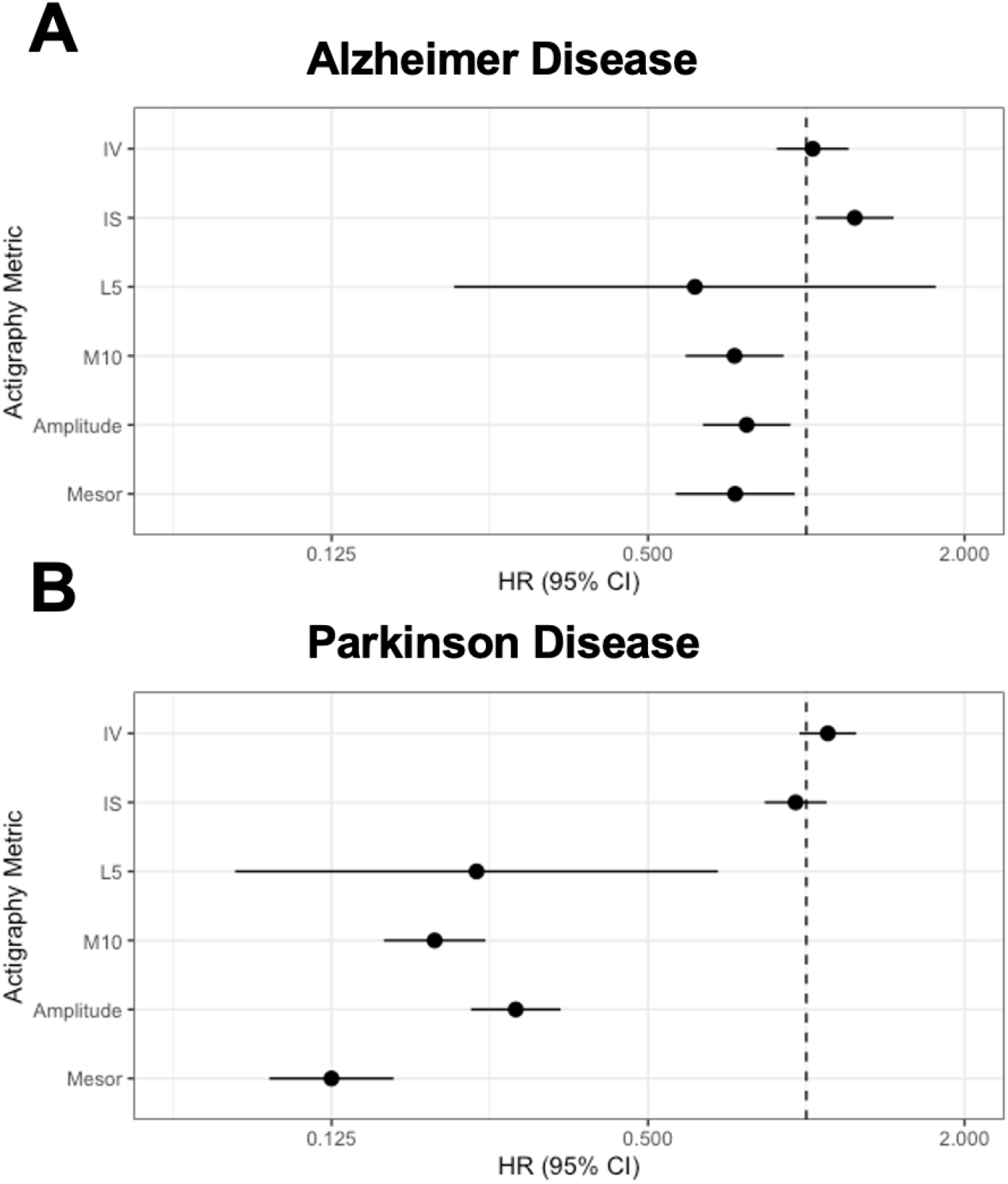
Adjusted hazard ratios for Alzheimer’s and Parkinson’s disease by 24-hour rhythms. Forest plots showing standardized hazard ratios (HR) per standard deviation increase and 95% confidence intervals (CI) for developing A) Alzheimer’s disease and B) Parkinson’s disease, from Cox regression models including age at actigraphy collection, gender, college education, baseline general health, baseline body mass index, and baseline Townsend deprivation index. Abbreviations: IV, intradaily variability; IS, interdaily stability, L5, least active 5 hours; M10, most active 10 hours.

Higher interdaily stability was associated with an increased risk of AD (hazard ratio [HR] per SD increase 1.24, 95% confidence interval [CI] 1.04-1.47). Lower M10 (HR 0.73, CI 0.59-0.91), amplitude (HR 0.77, CI 0.64-0.93), and mesor (HR 0.73, CI 0.56-0.95) were each associated with an increased risk of AD.

Lower L5 (HR 0.24, CI 0.08-0.68), M10 (HR 0.20, CI 0.16-0.25), amplitude (HR 0.28, CI 0.23-0.34), and mesor (HR 0.12, CI 0.10-0.16) were all associated with an increased risk of PD.

### fPCA

A visualization of mean 24-hour activity patterns across disease converters and matched controls is presented in **Figure 3**. Demographic information for individuals who developed AD and PD and the matched fPCA samples is presented in **Supplement Table 4**. The shapes and variance explained by the components were similar across AD and PD analyses and are presented in **Supplement Figures 2 and 3**. Component 1 could be described as an amplitude component, with higher values reflecting higher daytime activity, and explained 60% of the variance for the AD analysis and 64% for the PD analysis. Elevated component 2 represents later wake and bedtimes, with mainly flat peak activity, while lower component 2 represent earlier wake and bedtimes with a morning peak of activity; 16% of variance was captured in AD and 14% in PD analysis. Elevated component 3 represents a longer activity period interrupted by a midday dip in activity, possibly representing napping, while lower component 3 represents a flatter activity profile with slightly elevated morning activity; 11% of variance was captured in AD and 9.0% in PD analysis. Component 4, explaining only 6.8% for AD and 6.3% for PD, represents a late afternoon peak for those with low scores and a morning and evening peak for those with high scores.

**Figure 3.**
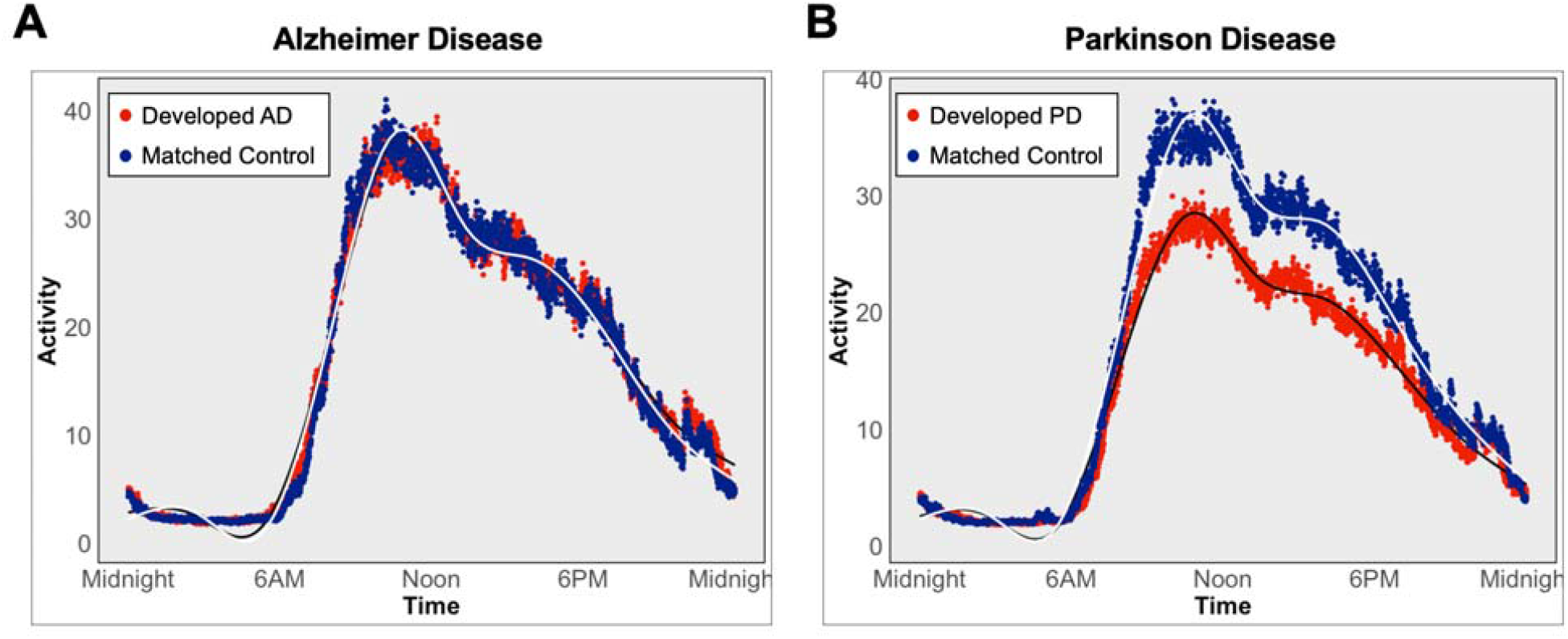
Average 24-hour activity curves for individuals who developed Alzheimer’s and Parkinson’s disease and matched individuals. Average activity curves in individuals who developed (A) Alzheimer’s disease or (B) Parkinson’s disease (red) or matched individuals who did not develop disease. Each plotted point represents a 30-second epoch of average activity. Daytime activity levels are visibly lower in individuals who developed Parkinson’s disease.

When magnitude scores for the four fPCA components were compared between individuals who converted to AD and matched controls, there were no differences between groups. In contrast, individuals who developed PD showed severely reduced component 1 scores relative to individuals who did not develop PD, suggesting lower daytime activity levels relative to matched controls (PD, -120.6 [419.9]; controls, 120.6 [411.5]; p<0.001). There were no differences for fPCA components 2, 3, or 4.

### Cognitive decline

Associations between rest-activity metrics and cognitive decline are presented in **Supplement Table 5** and visualized by quartiles in **Figure 4** and **Supplement Figure 4**. Worse longitudinal performance on the Symbol Digit Substitution Test was associated with lower mesor (β=0.004 ± 0.001, p=0.01), lower M10 (β=0.002 ± 0.001, p=0.01), and lower amplitude (β=0.002 ± 0.001, p=0.047). Increasing duration on Trails A, indicating worsening performance longitudinally, was associated with greater baseline intradaily variability (β=0.50 ± 0.15, p<0.001), lower interdaily stability (β=-0.89 ± 0.32, p=0.005), lower M10 (β=-0.005 ± 0.004, p=0.03), and lower amplitude (β=-0.012 ± 0.004, p=0.005). Increasing Trails B-A duration was associated with lower intradaily variability (β=-0.75 ± 0.24, p=0.001). Declining Numeric Memory performance was associated with greater intradaily variability (β=0.004 ± 0.002, p=0.01). Longitudinal change in fluid intelligence score was not associated with any baseline 24-hour rhythm metric.

**Figure 4.**
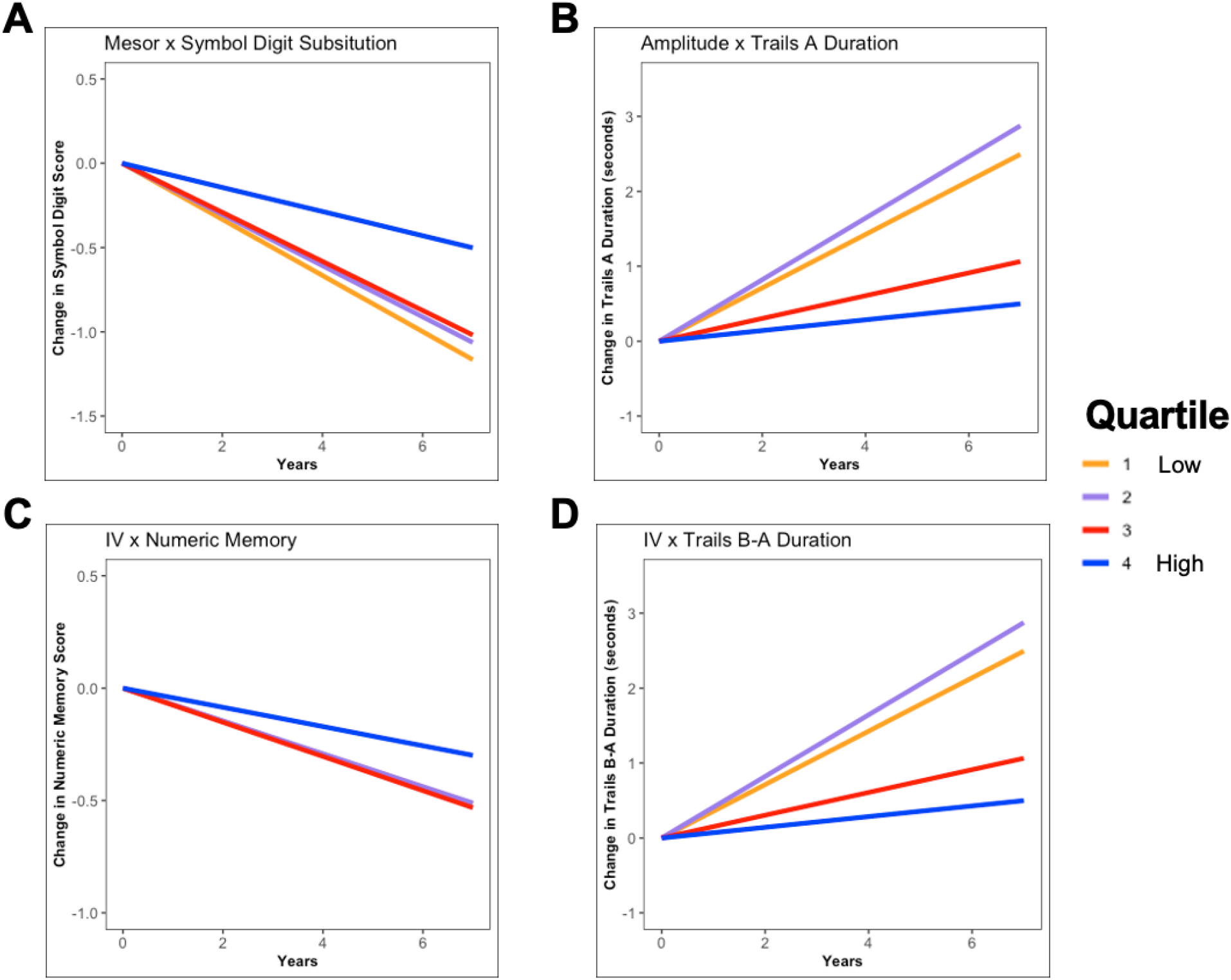
24-hour rhythms and longitudinal cognitive decline. Each plot represents beta estimates from a linear mixed effect model with a 24-hour rhythm metric as a predictor and cognitive performance as an outcome. Actigraphy metrics are split into quartiles for the purpose of visualizing associations with longitudinal change in cognitive test performance. Intercept is displayed as zero for all quartiles to visualize differences in slope between groups. (A) Higher mean activity (mesor) at baseline is associated with less decline on Symbol Digit test. (B) Greater amplitude at baseline is associated with better (faster) longitudinal performance on Trails A test. (C) Higher intradaily variability (IV) is associated with less decline on Numeric Memory test. (D) Higher IV is associated with better (faster) longitudinal performance on Trails B-A. All linear mixed models controlled for age at actigraphy collection, gender, college education, baseline general health, baseline body mass index, baseline Townsend deprivation index, and a binary variable indicating whether an individual’s baseline assessment was in-person or remote.

## DISCUSSION

In 82,829 older individuals with up to 8 years of follow-up, we found multiple metrics of actigraphy-measured 24-hour activity were associated with risk of developing Alzheimer’s disease, Parkinson’s disease, and subsequent cognitive decline. Reduced diurnal amplitude and activity levels were associated with a greater risk of both AD and PD, as well as greater declines in performance on the Symbol Digit and Trails A tests. Higher regularity of rhythms across days, (higher IS), was associated with a greater risk of AD, yet lower IS was associated with declining performance on Trails A. Lower activity during the least active five hours (L5) was associated with a higher risk of PD. A complementary data-driven approach (fPCA) revealed that individuals who converted to PD had significantly lower values for a daytime activity component (which explained 64% of the variance) compared to matched individuals who did not develop PD. Together, these data suggest objective measures of 24-hour activity could be used as community-based biomarkers of neurodegeneration risk.

Lower baseline activity levels (amplitude, mesor, and M10) were associated with a higher risk of both AD and PD conversion. The magnitude of these effects was greater for PD, with the majority of individuals who converted to PD being in the lowest quartile for amplitude, mesor, and M10. These results align with previous reports in smaller cohorts demonstrating links between reduced activity and diurnal amplitude and risk of dementia and cognitive impairment^13,14^ and PD^15^. Moreover, the matched fPCA analysis, which was naïve to the shape of 24-hour activity, revealed a daytime activity component that was lower in individuals who converted to PD, in agreement with the cosine-based analyses. This fPCA result suggests that daytime activity may be the most robust disease-specific difference in 24-hour activity that specifically determines those at risk for PD. Indeed, the fPCA in AD converters did not reveal differences in any of the 24-hour components, suggesting there was no consistent 24-hour activity “signature” that differed between those who converted to AD and matched non-converters.

Beyond activity and amplitude, greater regularity of rhythms across days (IS) was associated with a higher risk of conversion to AD. While this finding goes against a framework wherein circadian rhythms weaken in preclinical AD and are responsible in part for rhythm regularity^6,34^, it may reflect IS values being driven by factors other than circadian biology – for example, aging individuals with monotonous routines could be at increased risk for AD.

Multiple mechanisms may contribute to the link between reduced 24-hour rhythms and risk of incident PD. Brainstem circuits involved in sleep-wake regulation are affected early in the disease process, before the onset of dopaminergic loss ^3,34,36^. Relatedly, daytime napping^37^ and excessive daytime sleepiness^38,39^ are both associated with elevated risk of PD. Another explanation is that the presence of subclinical rigidity and bradykinesia in the prodromal phase of the disease may constrain movement, resulting in lower overall activity quantification^40^. This explanation is supported by our finding that PD risk was associated with lower L5 values, a proxy for nocturnal activity levels. Low L5 values in these individuals, rather than reflecting consolidated and restful sleep, are likely capturing nocturnal rigidity in prodromal disease stages. Importantly, low L5 was the only marker in our analysis specific in predicting subsequent PD diagnosis. Future studies with more detailed measures of sleep, motor impairment, and disease biomarkers could untangle the relative contributions of rigidity and impaired sleep-wake regulation to the observed associations.

The links between impaired 24-hour rhythms and subsequent cognitive decline are consistent with previous reports in smaller longitudinal samples of older adults, which have similarly reported lower mesor and amplitude predicting cognitive decline in women^10^ and in men^12^. In the present study, baseline amplitude, mesor, and M10 predicted worse longitudinal performance on the Symbol Digit Substitution Test, an assessment of processing speed and executive function^31^. Performance on Trails A, the numeric portion of the Trail Making Test and considered a measure of processing speed^32^, also showed greater decrease over time in individuals with lower baseline amplitude and M10. Higher amplitude, mesor, and M10 were also associated with a lower risk of developing AD. Together these associations agree with literature suggesting higher activity levels may contribute to preserved cognitive function^8,33^, or conversely that attenuated physical activity may serve as an indicator of looming decline.

The present associations between cognition and nonparametric measures of 24-hour rhythms offer a less straightforward interpretation. On one hand, declining performance on Trails A was associated with greater within-day rhythm fragmentation (IV) and weaker across-day rhythm stability (IS) at baseline. This finding is in line with hypothesized relationships between circadian rhythm robustness and cognitive aging^34,35^ as well as a recent report that high IV and low IS are associated with risk of cognitive impairment^11^. Conversely, we found higher IV was associated with *better* longitudinal performance on Trails B-A (a measure of executive task-switching) and Numeric Memory (a test of working memory), suggesting that individuals with more fragmented rhythms at baseline had less decline in these domains. These relationships raise the possibility that despite including multiple demographic and lifestyle covariates in our analyses, some cohort-specific effect may be driving IV values such that higher “fragmentation” of 24-hour activity actually reflects better brain health in at least a proportion of individuals.

## Limitations

This study has some limitations. First, the study did not include biological markers of disease or genetic information that could elucidate whether some individuals were at prodromal disease stages at the time of actigraphy collection. Second, we did not have objective sleep measures.

Actigraphy lacks specificity in differentiating sedentary activity from nocturnal sleep and daytime napping^41^. Given the absence of concurrent sleep logs or sleep timing information in the UK Biobank actigraphy data, we elected to take an agnostic approach by characterizing 24-hour sleep-wake rhythms rather than purporting to detect sleep based on activity. Finally, our study relied on ICD-10 codes and self-reported diagnoses to determine AD and PD, which could have led to underestimation of associations based on disease misclassifications.

## Conclusions

The present results suggest that 24-hour rhythm integrity as assessed by seven days of wrist actigraphy can serve as a prospective marker of incident AD and PD risk as well as cognitive decline.

## Data Availability

All data in the present study are available from the UK Biobank.

https://www.ukbiobank.ac.uk/

