## Supplement Figures and Tables for "Association of 24-hour activity patterns with risk of Alzheimer’s disease, Parkinson’s disease, and cognitive decline"

**Supplementary Online Content**

**Figure 1. CONSORT diagram of study inclusion.**

**Figure 2. Results of the functional principal components analysis for Alzheimer disease.**

**Figure 3.** **Results of the functional principal components analysis for Parkinson disease.**

**Figure 4. 24-hour rhythms and longitudinal change in all cognitive tests.**

**Table 1. Sample for longitudinal cognitive test data.**

**Table 2. Correlations between accelerometer-derived cosinor and nonparametric measures.**

**Table 3. Summary of survival analysis of 24-hour rhythms and developing Alzheimer and Parkinson disease.**

**Table 4. Demographics (at time of actigraphy collection) of participants who developed Alzheimer and Parkinson disease and matched controls in functional principal component analysis.**

**Table 5. Summary of 24-hour rhythm associations with longitudinal cognitive test performance.**

**Figure 1. CONSORT diagram of study inclusion.**

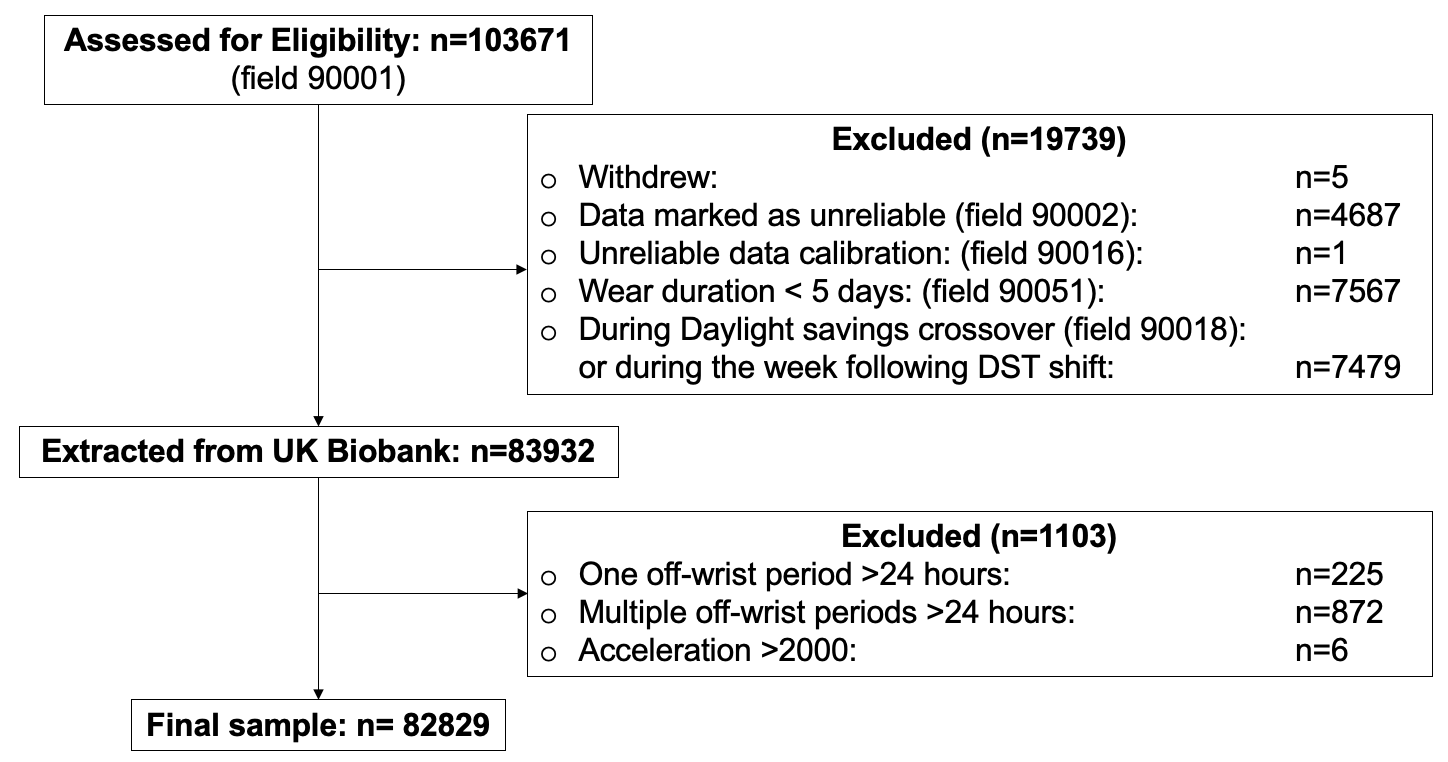

UK Biobank actigraphy data were collected from 103,670 total individuals. Individuals were excluded from the analytic sample if they withdrew from the study, had accelerometer data marked as unreliable, had unreliable calibration data, wore the accelerometer for less than five days, had their data collected during a daylight savings time crossover or the week following a daylight savings time shift, had at least one off-wrist period 24-hours or longer, or had implausibly high acceleration values (>2000). The remaining accelerometer data sample comprised 82,829 individuals.

**Figure 2. Results of the functional principal components analysis for Alzheimer disease.**

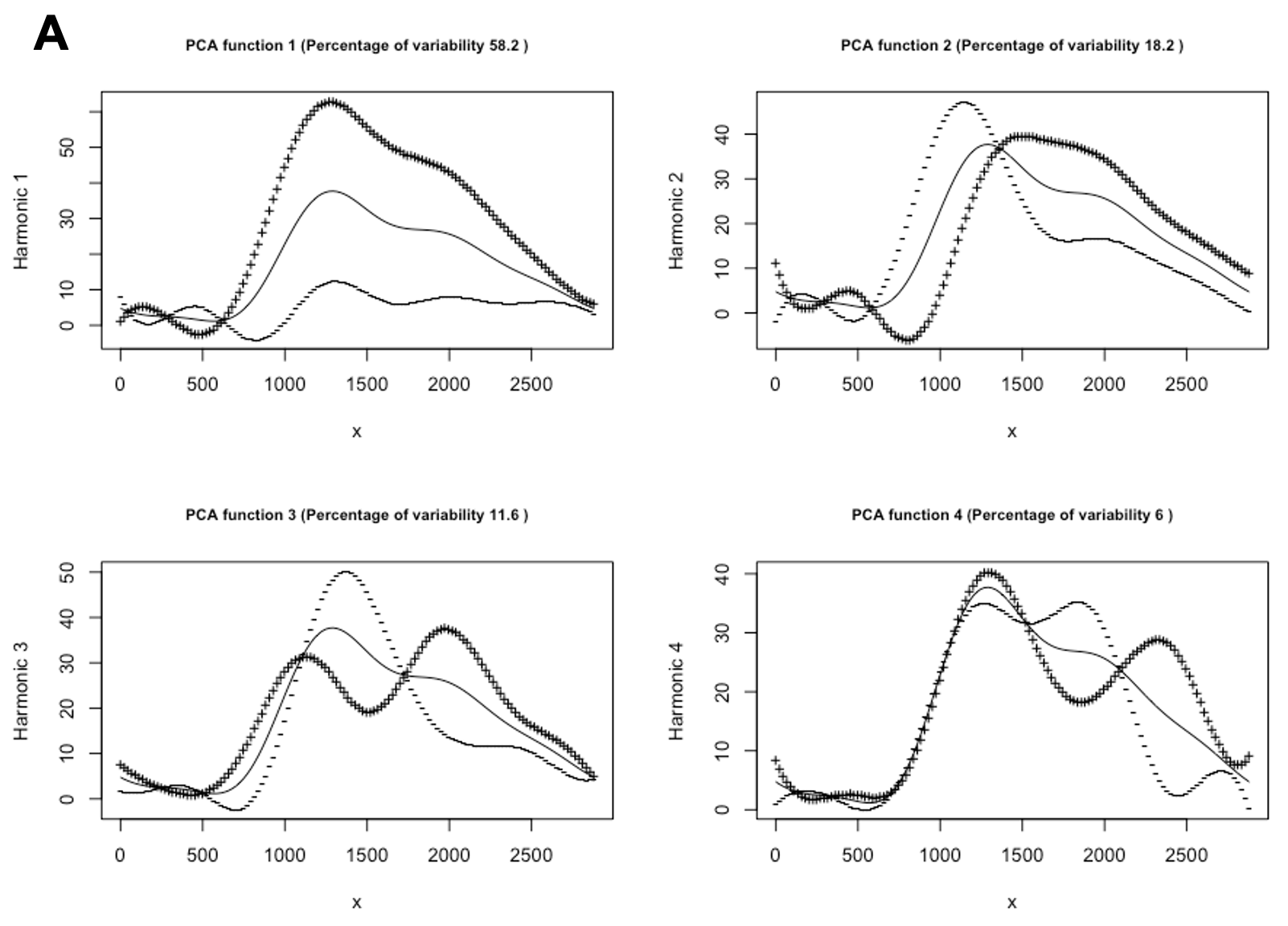

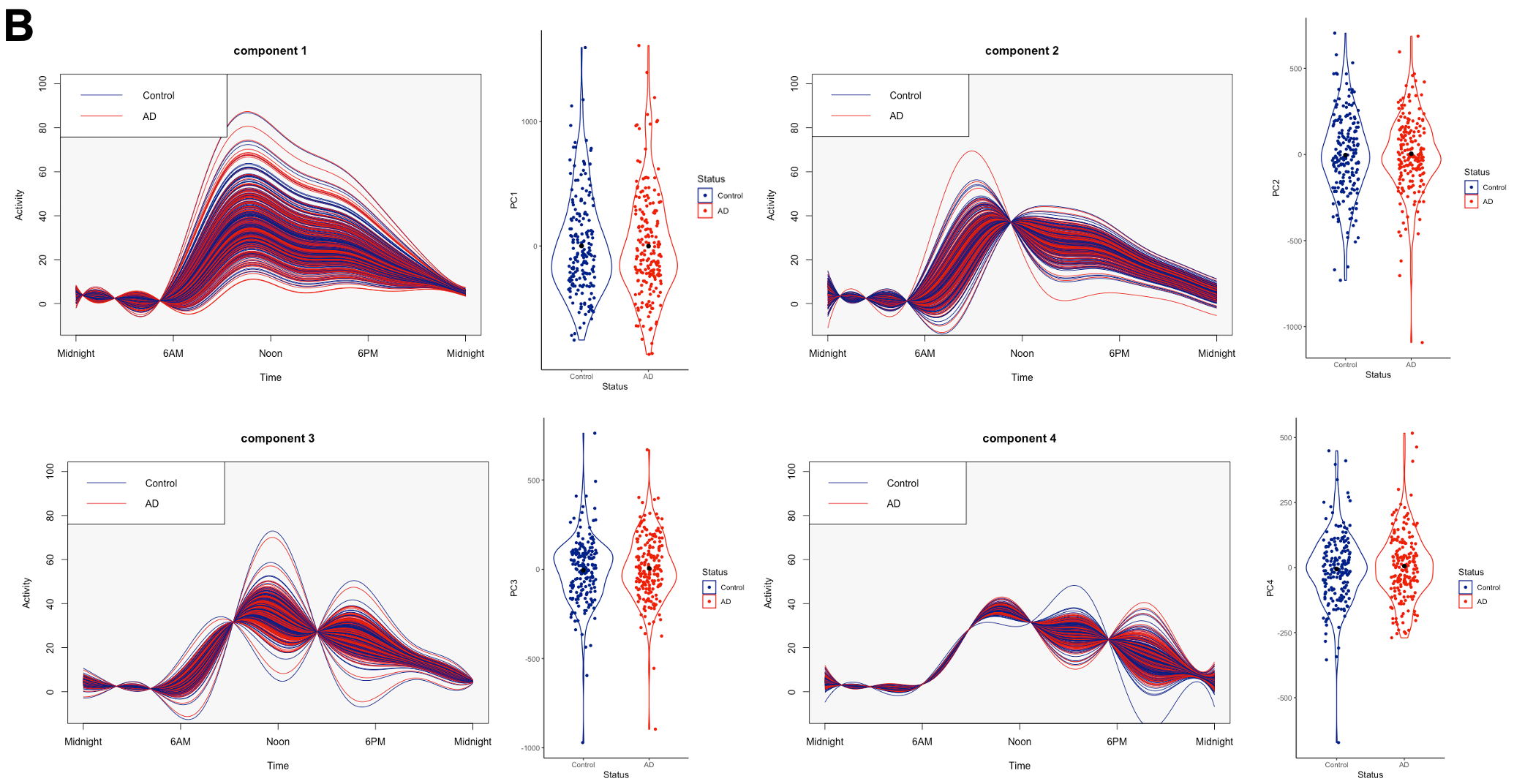

Four components were derived from 24-hour data from individuals who developed Alzheimer disease and matched controls who did not develop Alzheimer disease. (A) The solid line represents the same average activity curve in each of the four plots. Each of the four components is visualized by adding (+) or subtracting (-) the component to the average activity curve. (B) For each of the four components, the left plot shows individual curves for individuals who developed Alzheimer disease (AD) and matched individuals who did not (Control). The right violin plot shows weights for every individual with a black dot representing the group mean.

**Figure 3. Results of the functional principal components analysis for Parkinson disease.**

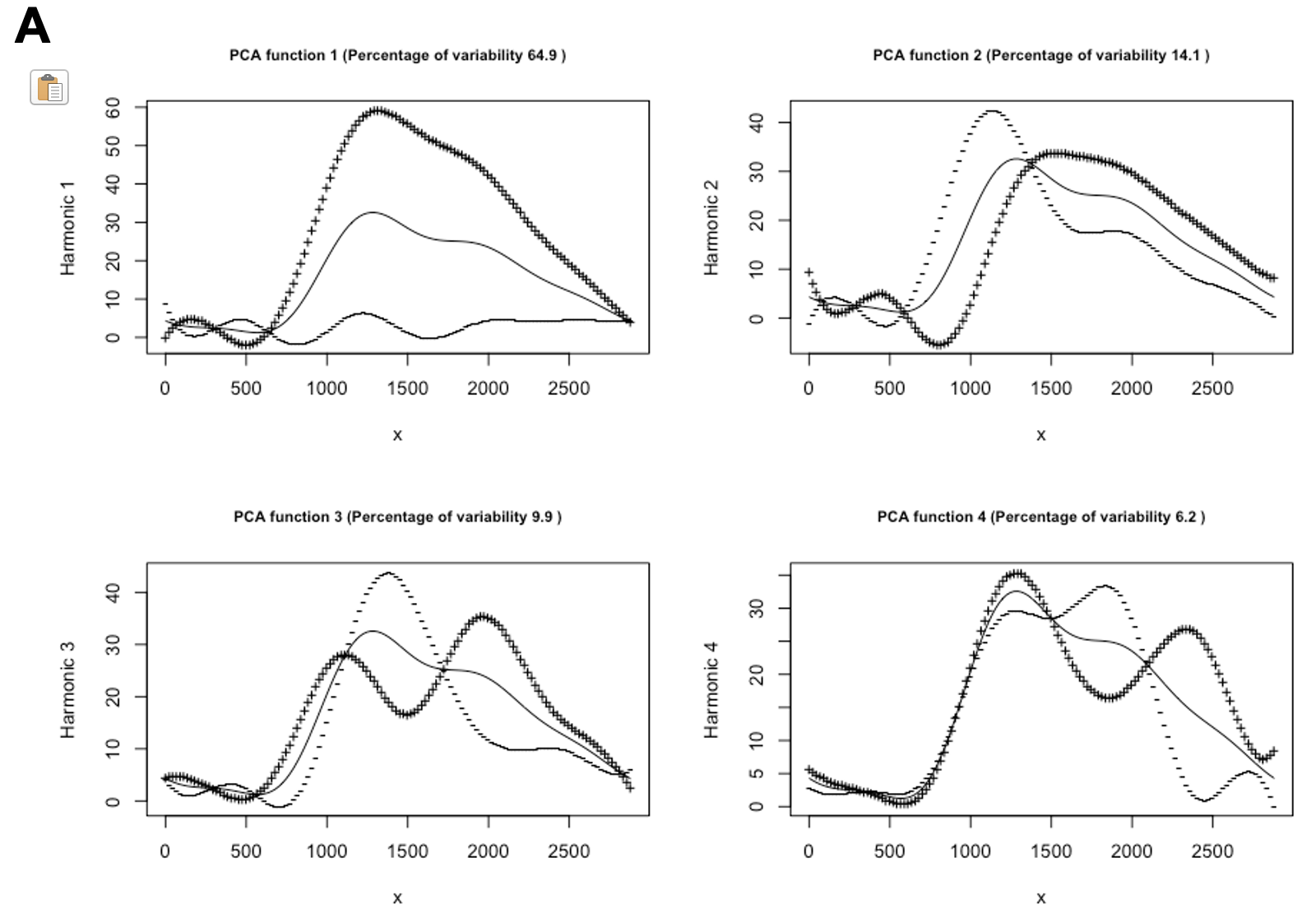

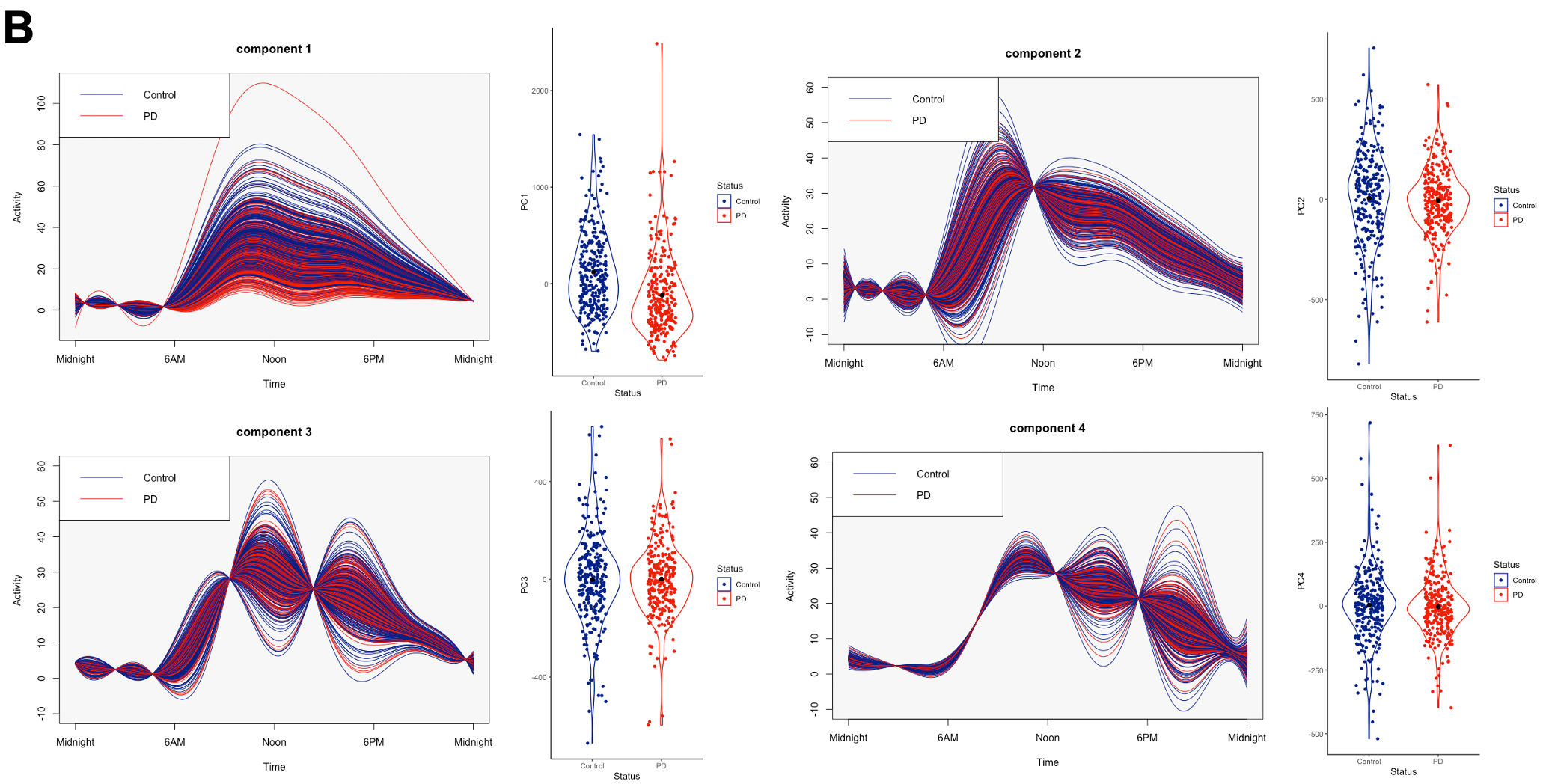

Four components were derived from 24-hour data from individuals who developed Parkinson disease and matched controls who did not develop Parkinson disease. (A) The solid line represents the same average activity curve in each of the four plots. Each of the four components is visualized by adding (+) or subtracting (-) the component to the average activity curve. (B) For each of the four components, the left plot shows individual curves for individuals who developed Parkinson disease (PD) and matched individuals who did not (Control). The right violin plot shows weights for every individual with a black dot representing the group mean.

**Figure 4. 24-hour rhythms and longitudinal change in all cognitive tests.**

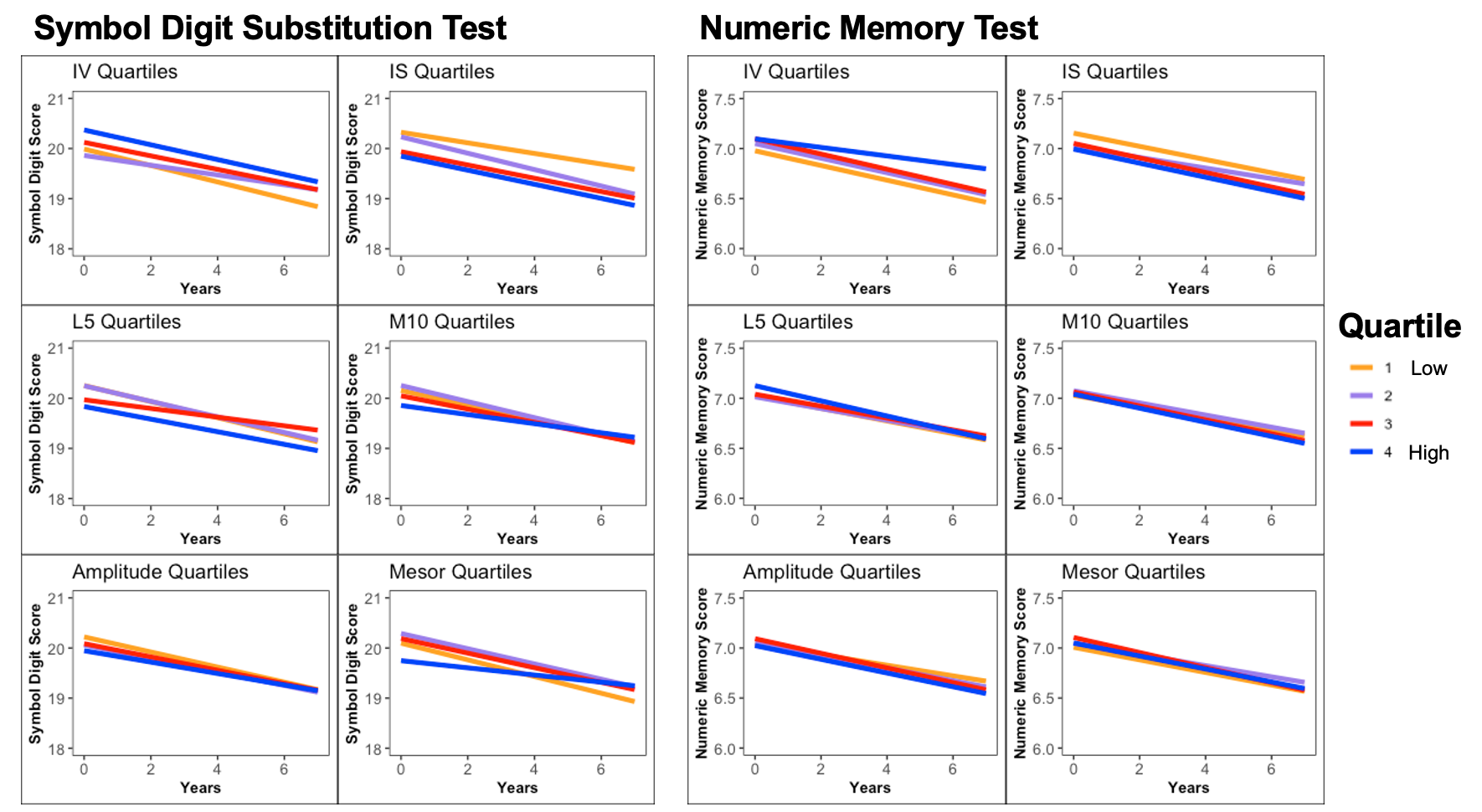

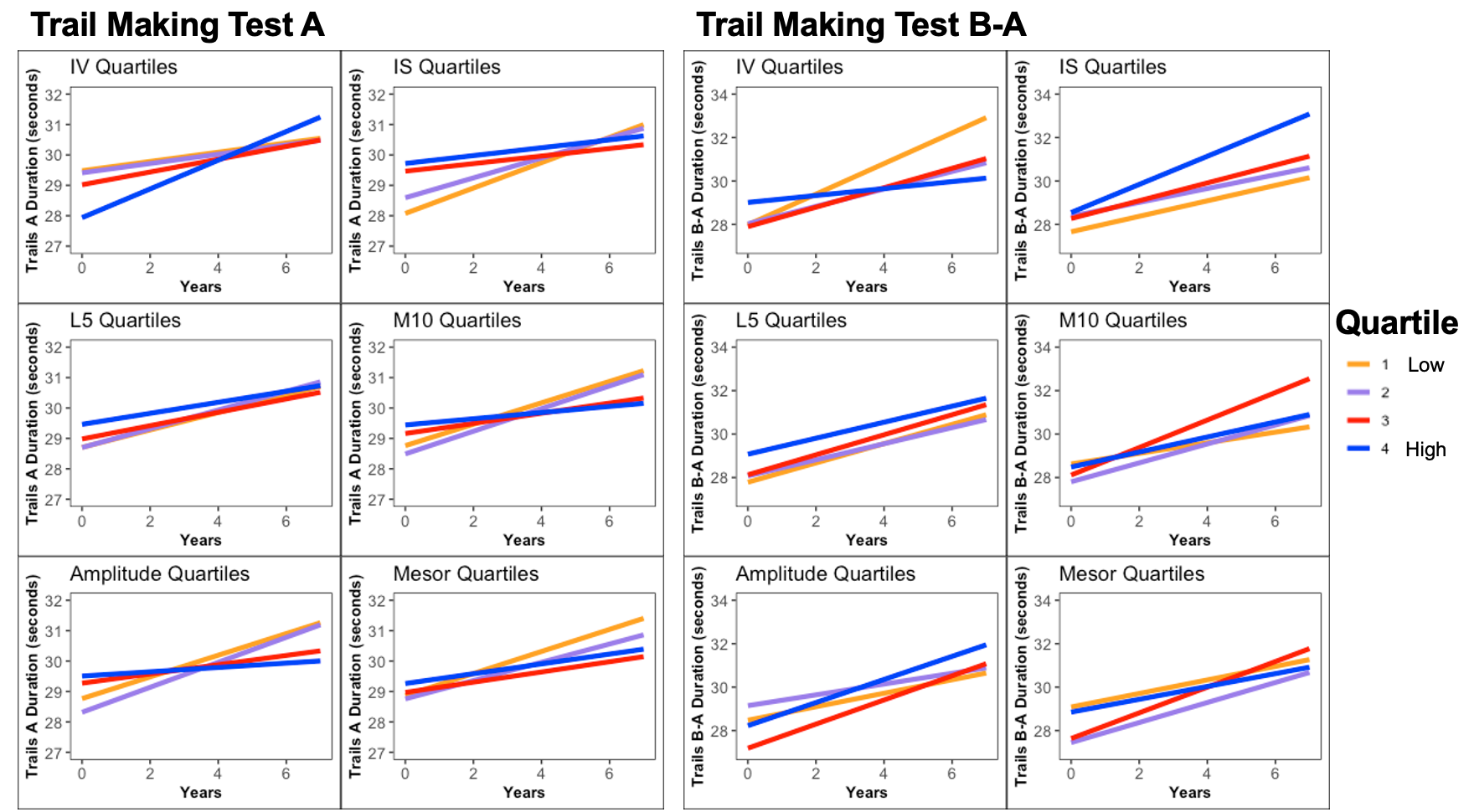

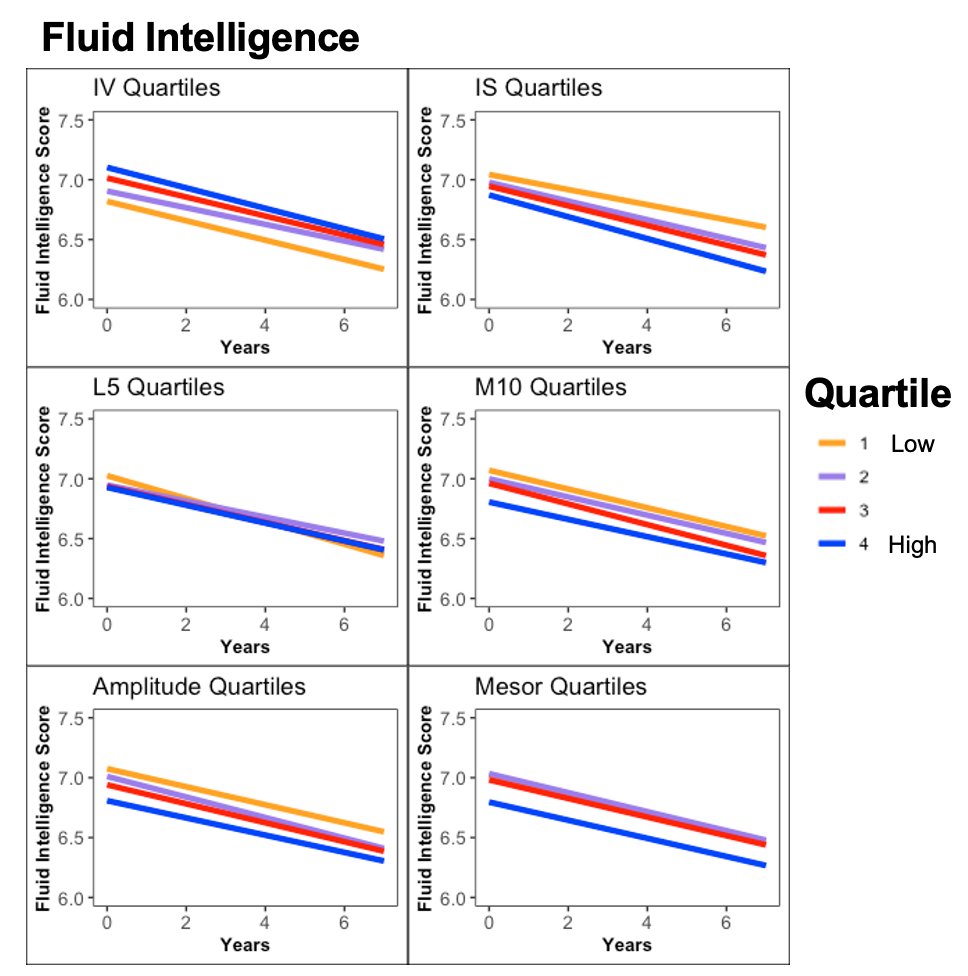

Each plot represents beta estimates from a linear mixed model. All linear mixed models controlled for age at actigraphy collection, gender, college education, baseline general health, baseline body mass index, and baseline Townsend deprivation index. All linear mixed models included a random intercept for each participant, and an interaction with time for the actigraphy measure of interest and each covariate. Models also included a binary variable indicating whether an individual’s baseline assessment was in-person or remote. Abbreviations: IV, intradaily variability; IS, interdaily stability, L5, least active 5 hours; M10, most active 10 hours.

**Table 1. Sample for longitudinal cognitive test data.**

|  | **Symbol Digit** | **Trails A** | **Trails B-A** | **Numeric Memory** | **Fluid Intelligence** |
| --- | --- | --- | --- | --- | --- |
| **Longitudinal N** | 7,533 | 6,731 | 6,629 | 7,401 | 11,030 |
| **# time points** | 2.1 ± 0.3 | 2.1 ± 0.3 | 2.1 ± 0.3 | 2.1 ± 0.3 | 2.1 ± 0.3 |
| **Follow-up years** | 4.3 ± 1.1 | 4.3 ± 1.1 | 4.3 ± 1.1 | 4.3 ± 1.1 | 3.3 ± 1.9 |

Number of individuals included in longitudinal analyses for each cognitive test. Mean ± standard deviation is listed for number time points and years of follow-up.

**Table 2. Correlations between accelerometer-derived cosinor and nonparametric measures.**

| **IV** | 1 |  |  |  |  |  |
| --- | --- | --- | --- | --- | --- | --- |
| **IS** | -0.45 | 1 |  |  |  |  |
| **L5** | 0.00 | -0.06 | 1 |  |  |  |
| **M10** | -0.12 | 0.03 | 0.47 | 1 |  |  |
| **Amplitude** | -0.21 | 0.10 | 0.01 | 0.89 | 1 |  |
| **Mesor** | -0.06 | -0.05 | 0.70 | 0.93 | 0.70 | 1 |
|  | **IV** | **IS** | **L5** | **M10** | **Amplitude** | **Mesor** |

Bivariate correlations (r value) between actigraphy measures across all participants. Abbreviations: IV, intradaily variability; IS, interdaily stability, L5, least active 5 hours; M10, most active 10 hours.

**Table 3. Summary of survival analysis of 24-hour rhythms and developing Alzheimer and Parkinson disease.**

|  | **Alzheimer Disease** (n=191) | | | **Parkinson Disease** (n=266) | | |
| --- | --- | --- | --- | --- | --- | --- |
|  | *HR* | *95% CI* | *p-value* | *HR* | *95% CI* | *p-value* |
| **IV** | 1.03 | 0.88-1.20 | 0.73 | 1.10 | 0.97-1.24 | 0.14 |
| **IS** | 1.24 | 1.04-1.47 | 0.01 | 0.95 | 0.83-1.09 | 0.50 |
| **L5** | 0.61 | 0.21-1.76 | 0.37 | 0.24 | 0.08-0.68 | 0.01 |
| **M10** | 0.73 | 0.59-0.91 | 0.004 | 0.20 | 0.16-0.25 | <0.001 |
| **Amplitude** | 0.77 | 0.64-0.93 | 0.01 | 0.28 | 0.23-0.34 | <0.001 |
| **Mesor** | 0.73 | 0.56-0.95 | 0.02 | 0.12 | 0.1-0.16 | <0.001 |

Abbreviations: HR, hazard ratio; CI, confidence interval, IV, intradaily variability; IS, interdaily stability, L5, least active 5 hours; M10, most active 10 hours. All Cox proportional hazards regression models controlled for age at actigraphy collection, gender, college education, baseline general health, baseline body mass index, and baseline Townsend deprivation index. See Figure 3 for visualization of these data.

**Table 4. Demographics (at time of actigraphy collection) of participants who developed Alzheimer and Parkinson disease and matched controls in functional principal component analysis.**

|  | **Alzheimer Disease** | | **Parkinson Disease** | |
| --- | --- | --- | --- | --- |
|  | **Cases** (n=191) | **Controls**  (n=191) | **Cases**  (n=266) | **Controls**  (n=266) |
| **Age** | 69.9 ± 4.5 | 69.8 ± 4.5 | 68.6 ± 5.1 | 68.7 ± 5.2 |
| **Female N (%)** | 100 (52) | 97 (51) | 86 (32) | 86 (32) |
| **General Health [1-4]** | 2.1 ± 0.7 | 2.1 ± 0.6 | 2.1 ± 0.7 | 2.1 ± 0.6 |
| **College Education N (%)** | 64 (34) | 61 (32) | 109 (41) | 109 (41) |
| **Body mass index** | 26.7 ± 4.6 | 26.9 ± 4.3 | 26.8 ± 4.3 | 26.9 ± 4.0 |
| **Townsend Deprivation Index** | -1.7 ± 2.9 | -1.6 ± 3.0 | -2.1 ± 2.6 | -1.9 ± 2.7 |
| **Years to Diagnosis** | 4.8 ± 1.7 | NA | 4.2 ± 1.9 | NA |

**Table 5. Summary of 24-hour rhythm associations with longitudinal cognitive test performance.**

|  | **Symbol Digit**  (n=7,533) | | **Trails A**  (n=6,731) | | **Trails B-A**  (n=6,629) | | **Numeric Memory**  (n=7,401) | | **Fluid Intelligence**  (n=11,030) | |
| --- | --- | --- | --- | --- | --- | --- | --- | --- | --- | --- |
|  | *β (SE)* | *p-value* | *β (SE)* | *p-value* | *β (SE)* | *p-value* | *β (SE)* | *p-value* | *β (SE)* | *p-value* |
| **IV*time** | -0.005 (0.05) | 0.92 | 0.50 (0.15) | 0.001 | -0.75 (0.24) | 0.001 | 0.05 (0.02) | 0.01 | -0.01 (0.02) | 0.42 |
| **IS*time** | -0.10 (0.10) | 0.34 | -0.89 (0.32) | 0.005 | 0.51 (0.51) | 0.32 | -0.06 (0.04) | 0.12 | -0.07 (0.04) | 0.05 |
| **L5*time** | 0.01 (0.01) | 0.38 | -0.02 (0.03) | 0.41 | -0.01 (0.04) | 0.86 | 0.001 (0.003) | 0.72 | 0.003 (0.003) | 0.39 |
| **M10*time** | 0.002 (0.001) | 0.01 | -0.01 (0.002) | 0.03 | 0.001 (0.004) | 0.80 | -0.0002 (0.0003) | 0.59 | 0.0002 (0.0003) | 0.51 |
| **Amplitude*time** | 0.002 (0.001) | 0.048 | -0.01 (0.004) | 0.005 | 0.004 (0.01) | 0.52 | -0.0003 (0.0004) | 0.49 | 0.0003 (0.0005) | 0.47 |
| **Mesor*time** | 0.004 (0.001) | 0.01 | -0.01 (0.004) | 0.13 | -0.003 (0.01) | 0.63 | -0.0003 (0.0005) | 0.59 | 0.0005 (0.001) | 0.36 |

Abbreviations: IV, intradaily variability; IS, interdaily stability, L5, least active 5 hours; M10, most active 10 hours. All linear mixed-effects models controlled for age at actigraphy collection, gender, college education, baseline general health, baseline body mass index, and baseline Townsend deprivation index. All linear mixed models included a random intercept for each participant, and an interaction with time for the actigraphy measure of interest and each covariate. Models also included a binary variable indicating whether an individual’s baseline assessment was in-person or remote.
